# Estimated Free-Living Energy Balance and Perceived Physical and Psychological Stress in Japanese Male University Students: A 28-Day Observational Study

**DOI:** 10.64898/2026.10.13.26365271

**Authors:** Asumi Yoshida

## Abstract

**Background:** Whether whole-body energy balance is associated with perceived stress in young adults is unclear.

**Aim:** To examine associations of estimated free-living energy balance with physical and psychological stress during 28-day monitoring.

**Methods:** This secondary analysis of an observational study included 18 Japanese male university students with at least 14 valid accelerometer days (at least 600 min/day). Energy intake was estimated using the Brief-type Self-administered Diet History Questionnaire; total energy expenditure was estimated from accelerometry plus resting metabolic rate. Physical and psychological stress were assessed before and after monitoring using 100-mm visual analog scales. Baseline-adjusted linear regression was used.

**Summary:** Mean energy balance was −79 ± 531 kcal/day. Energy balance was not clearly associated with post-monitoring physical stress (B = 0.067 per 100 kcal/day, 95% CI −1.604 to 1.737; *p* = 0.933) or psychological stress (B = 0.314, 95% CI −2.571 to 3.199; *p* = 0.820). No clear association between estimated free-living energy balance and post-monitoring stress was detected.

## Introduction

Energy balance reflects the difference between energy intake and total energy expenditure and may influence fatigue, recovery, and well-being. Evidence linking inadequate energy status with psychological outcomes has largely arisen from athletic populations and the literature on low energy availability and Relative Energy Deficiency in Sport (REDs) [1, 2]. However, energy availability and whole-body energy balance are distinct constructs, and evidence relating free-living whole-body energy balance to perceived stress in healthy young adults remains limited.

University students are relevant because dietary intake, physical activity, and sleep may vary with academic and social schedules. Physical activity is often associated with lower perceived stress in university students, although findings are heterogeneous [3], while poor sleep and insomnia symptoms are associated with greater stress [4]. These behaviors may therefore provide useful secondary context when examining energy balance and stress.

This secondary analysis examined healthy Japanese male university students during 28 days of free-living monitoring. The primary aim was to determine whether estimated energy balance was associated with post-monitoring physical stress after adjustment for baseline physical stress. We hypothesized that a more negative energy balance would be associated with greater physical stress. Psychological stress, self-reported sleep period, moderate-to-vigorous physical activity (MVPA), and sedentary time were examined in secondary analyses.

## Methods

### Participants and design

This was a secondary analysis of a prospective observational study involving 28 days of free-living monitoring between August 2021 and July 2022. Twenty-nine healthy Japanese male university students were recruited through advertisements and direct announcements at a university in Japan. Eligibility included absence of diseases requiring a special diet or dietary restriction.

For the present analysis, participants were required to have at least 14 valid accelerometer days, with a valid day defined as at least 600 min/day of wear time. This standardized minimum wear-time criterion was adopted to ensure adequate daily coverage for characterizing habitual free-living activity and total energy expenditure and is consistent with previous HJA-750C research [5]. The analytic sample comprised all participants from the original cohort who met these criteria; no additional sampling was performed.

### Measurements

Body composition was assessed using multifrequency bioelectrical impedance (MC-780A-N; Tanita corp., Tokyo, Japan). Resting metabolic rate (RMR) was measured after a fasting period of at least 12 h by indirect calorimetry using Douglas bags and calculated using the Weir equation [6].

Total energy intake (TEI) was estimated using the Brief-type Self-administered Diet History Questionnaire (BDHQ), a 58-item fixed-portion-type questionnaire assessing habitual dietary intake during the previous month [7, 8]. Questionnaires were checked for unanswered items; participants were asked to complete or clarify omissions, when necessary, after which questionnaires were submitted to the BDHQ analysis center for standard processing. Energy and nutrient estimates were derived using the standard BDHQ algorithm based on the Standard Tables of Food Composition in Japan. Dietary supplements were not included in estimated TEI. No participant was excluded on the basis of the magnitude of reported TEI.

Total energy expenditure (TEE) was estimated from accelerometer-derived energy expenditure combined with individually measured RMR. The Active style Pro has demonstrated validity for estimating physical activity energy expenditure under free-living conditions against the doubly labeled water method [9], and approaches combining accelerometry with individually measured RMR have been used to estimate free-living TEE [10]. Estimated energy balance was calculated as TEI minus TEE; negative values indicated an estimated energy deficit. Because questionnaire-based assessment has limited validity for absolute energy intake, energy balance was treated as an estimated exposure, and no statistical correction for dietary measurement error was applied [11].

Physical activity was assessed using a waist-worn triaxial accelerometer (Active style Pro HJA-750C; Omron Healthcare Co., Ltd., Kyoto, Japan) in 60-s epochs. Participants wore the device during waking hours and recorded attachment and removal times in a daily diary. Sequences of at least 60 consecutive minutes with zero activity counts were reviewed and classified as non-wear. A valid day required at least 600 min of wear, and at least 14 valid days were required for analysis. Participant-level values were averaged across valid days. Physical activity intensity was estimated using the validated HJA-750C algorithm [12]. Activity was classified as sedentary (> 0 to ≤ 1.5 METs), light (> 1.5 to < 3.0 METs), moderate (≥ 3.0 to < 6.0 METs), or vigorous (≥ 6.0 METs); MVPA was the sum of moderate and vigorous activity [12]. For TEE estimation, periods without accelerometer-derived intensity data were assigned energy expenditure equivalent to individual RMR.

Self-reported sleep period was calculated from diary-recorded bedtime to wake time and did not represent objectively measured sleep duration. Physical and psychological stress were assessed immediately before and after monitoring using separate 100-mm visual analog scales (VAS), ranging from no stress to unbearable stress; higher scores indicated greater perceived stress [13]. Assessments were conducted at approximately the same time of day whenever possible.

### Statistical analysis

Analyses were performed using IBM SPSS Statistics version 28. Continuous variables are reported as mean ± SD. Pre-post changes in stress were assessed using paired-samples *t* tests with paired-samples Cohen’s *d*; Wilcoxon signed-rank tests were sensitivity analyses.

The primary linear regression model included post-monitoring physical stress as the outcome and baseline physical stress plus estimated energy balance as predictors. Baseline stress was included to account for pre-existing between-participant differences in stress. Energy balance was scaled per 100 kcal/day. The corresponding psychological-stress model was secondary. For transparency, unadjusted energy-balance models were also reported. Self-reported sleep period (per 1 h/day), MVPA (per 30 min/day), and sedentary time (per 1 h/day) were each examined in separate models adjusted for the corresponding baseline stress score. Results are reported as unstandardized coefficients (B), 95% confidence intervals (CI), standardized coefficients (*β*), and adjusted R^2^.

Residual plots, variance inflation factors, and Cook’s distance were examined. Fat-free mass was added to the primary model as a sensitivity analysis. An influence analysis was performed for the physical stress–sedentary time model because one observation had Cook’s distance > 1. No subgroup or interaction analyses were performed. Among analyzed participants, no data were missing for variables used in the reported analyses. All tests were two-sided with *p* < 0.05. Given the exploratory design, interpretation emphasized effect estimates and 95% CIs without formal multiplicity adjustment.

## Results

Of 29 enrolled participants, 18 met the current accelerometer criterion of at least 14 valid days with at least 600 min/day of wear and were analyzed; 11 did not meet this criterion. Among these 18 participants, dietary data were complete, and no exclusion based on implausible reported energy intake was applied. Participants contributed 22.4 ± 4.5 valid accelerometer days (range 14–28). Mean age was 20.4 ± 2.0 years, body mass index 23.0 ± 2.8 kg/m^2^, TEI 2,270 ± 850 kcal/day, TEE 2,378 ± 481 kcal/day, and estimated energy balance −79 ± 531 kcal/day (Table 1).

**Table 1.**
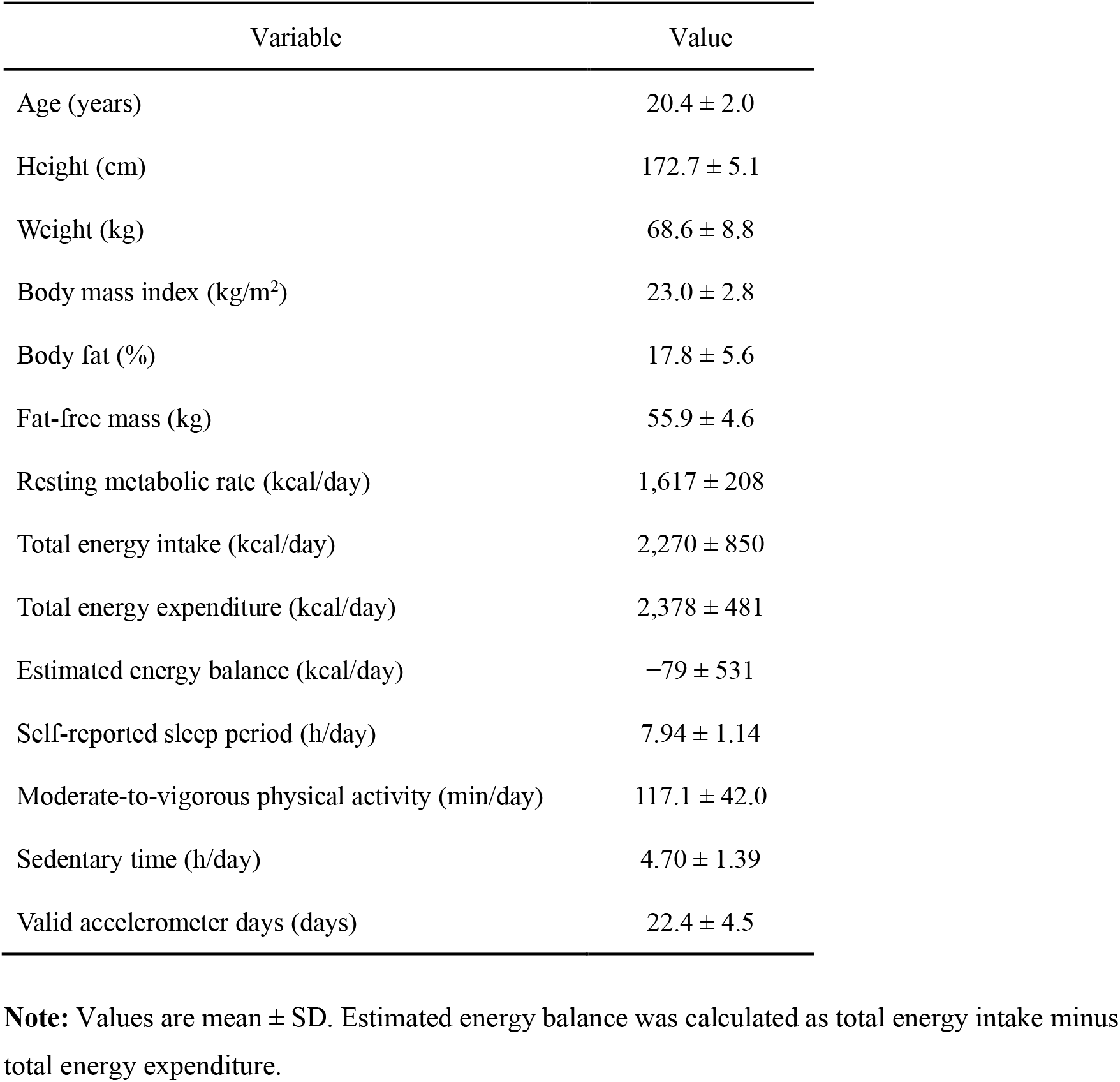
Participant characteristics (n = 18)

| Variable | Value |
| --- | --- |
| Age (years) | 20.4 ± 2.0 |
| Height (cm) | 172.7 ± 5.1 |
| Weight (kg) | 68.6 ± 8.8 |
| Body mass index (kg/m<sup>2</sup>) | 23.0 ± 2.8 |
| Body fat (%) | 17.8 ± 5.6 |
| Fat-free mass (kg) | 55.9 ± 4.6 |
| Resting metabolic rate (kcal/day) | 1,617 ± 208 |
| Total energy intake (kcal/day) | 2,270 ± 850 |
| Total energy expenditure (kcal/day) | 2,378 ± 481 |
| Estimated energy balance (kcal/day) | −79 ± 531 |
| Self-reported sleep period (h/day) | 7.94 ± 1.14 |
| Moderate-to-vigorous physical activity (min/day) | 117.1 ± 42.0 |
| Sedentary time (h/day) | 4.70 ± 1.39 |
| Valid accelerometer days (days) | 22.4 ± 4.5 |

**Note:** Values are mean ± SD. Estimated energy balance was calculated as total energy intake minus total energy expenditure.

Physical stress decreased from 31.78 ± 26.17 to 26.39 ± 22.68 (change −5.39, 95% CI −14.87 to 4.09; *p* = 0.247; *d* = −0.28). Psychological stress changed from 37.94 ± 24.79 to 40.50 ± 27.32 (change 2.56, 95% CI −10.46 to 15.57; *p* = 0.684; *d* = 0.10). Wilcoxon analyses led to the same conclusions. Individual trajectories varied substantially (Figure 1).

**Figure 1.**
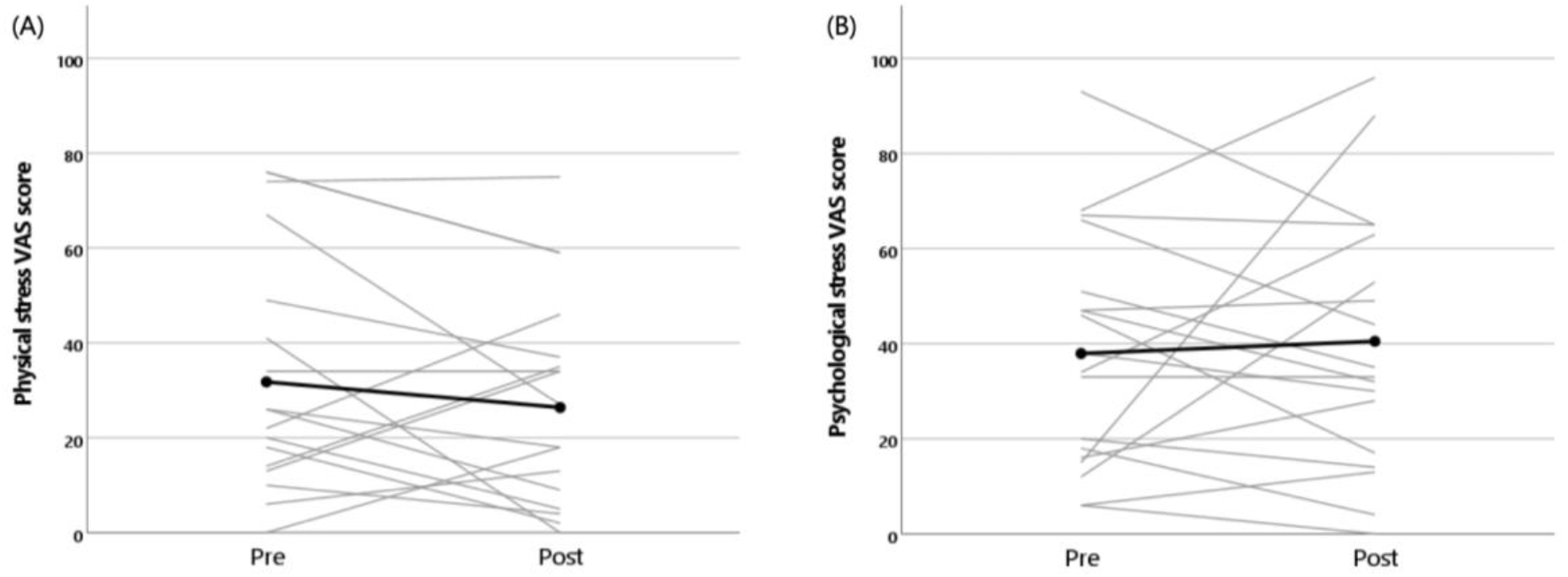
Individual changes in physical and psychological stress VAS scores from pre- to post-monitoring. **Note:** (A) Physical stress and (B) psychological stress. Gray lines represent individual participants (n = 18), and the thick black line with filled circles represents the group mean. VAS scores range from 0 to 100, with higher scores indicating greater perceived stress.

In unadjusted models, estimated energy balance was not clearly associated with post-monitoring physical stress (B = −0.057 per 100 kcal/day, 95% CI −2.321 to 2.206; *β* = −0.013; *p* = 0.958) or psychological stress (B = 1.585, 95% CI −1.009 to 4.179; *β* = 0.308; *p* = 0.214). After baseline adjustment, energy balance was still not clearly associated with physical stress (B = 0.067, 95% CI −1.604 to 1.737; *β* = 0.016; *p* = 0.933; adjusted R^2^=0.428) or psychological stress (B = 0.314, 95% CI −2.571 to 3.199; *β* = 0.061; *p* = 0.820; adjusted R^2^ = 0.152). Baseline physical stress was associated with post-monitoring physical stress (B = 0.611, 95% CI 0.272 to 0.950; *β* = 0.704; *p* = 0.002).

No clear baseline-adjusted associations were observed for self-reported sleep period, MVPA, or sedentary time with either stress outcome (Table 2). Adding fat-free mass to the primary model did not materially alter the energy-balance estimate (B = −0.145, 95% CI −1.944 to 1.654; *p* = 0.865). One observation was influential in the physical stress–sedentary time model (Cook’s distance = 1.326); after exclusion, the sedentary-time coefficient changed from −0.205 to −4.934 points/h but remained imprecise (95% CI −12.447 to 2.579; *p* = 0.181).

**Table 2.** Associations with post-monitoring physical and psychological stress VAS scores.

| Outcome | Exposure | Model | B | 95% CI | $\beta$ | *p* | Adjusted R<sup>2</sup> |
| --- | --- | --- | --- | --- | --- | --- | --- |
| Physical stress | Energy balance, per 100 kcal/day | Unadjusted | −0.057 | −2.321 to 2.206 | −0.013 | 0.958 | −0.062 |
|  | Energy balance, per 100 kcal/day | Baseline-adjusted | 0.067 | −1.604 to 1.737 | 0.016 | 0.933 | 0.428 |
|  | Sleep period, per 1 h/day | Baseline-adjusted | 0.155 | −8.703 to 9.014 | 0.008 | 0.971 | 0.428 |
|  | MVPA, per 30 min/day | Baseline-adjusted | −0.930 | −7.248 to 5.388 | −0.057 | 0.758 | 0.432 |
|  | Sedentary time, per 1 h/day | Baseline-adjusted | −0.205 | −6.781 to 6.371 | −0.013 | 0.948 | 0.428 |
| Psychological stress | Energy balance, per 100 kcal/day | Unadjusted | 1.585 | −1.009 to 4.179 | 0.308 | 0.214 | 0.038 |
|  | Energy balance, per 100 kcal/day | Baseline-adjusted | 0.314 | −2.571 to 3.199 | 0.061 | 0.820 | 0.152 |
|  | Sleep period, per 1 h/day | Baseline-adjusted | −4.037 | −15.997 to 7.923 | −0.168 | 0.483 | 0.178 |
|  | MVPA, per 30 min/day | Baseline-adjusted | −2.886 | −12.164 to 6.393 | −0.148 | 0.517 | 0.173 |
|  | Sedentary time, per 1 h/day | Baseline-adjusted | 5.546 | −3.340 to 14.432 | 0.283 | 0.203 | 0.239 |

**Note:** Unadjusted models contained energy balance only. Baseline-adjusted models additionally included the corresponding baseline physical or psychological stress VAS score. Each lifestyle exposure was examined in a separate model. Energy balance in the physical-stress analysis was the primary exposure; remaining analyses were secondary. B, unstandardized regression coefficient; $\beta$ , standardized regression coefficient; CI, confidence interval; MVPA, moderate-to-vigorous physical activity; VAS, visual analog scale.

## Discussion

In this 28-day exploratory study, estimated free-living energy balance was not clearly associated with subsequent physical or psychological stress after accounting for baseline stress. The hypothesized association between a more negative energy balance and greater physical stress was not supported. Baseline physical stress, rather than energy balance, was the strongest predictor of post-monitoring physical stress. Secondary analyses likewise showed no clear associations for self-reported sleep period, MVPA, or sedentary time.

These findings should not be interpreted as evidence that energy status is unrelated to stress. Evidence linking inadequate energy status with fatigue and psychological outcomes largely concerns low energy availability and REDs [1] [2], which differ conceptually from whole-body energy balance. Moreover, TEI was questionnaire-derived. Although the BDHQ has demonstrated reasonable validity for ranking many food-group and energy-adjusted nutrient intakes among Japanese adults [7] [8], estimation of absolute energy intake remains subject to substantial reporting error [11]. Error in TEI or TEE propagates into their difference and could obscure associations, especially in a small sample. The BDHQ represented approximately one month of habitual intake, broadly corresponding to monitoring, but did not capture day-to-day energy-balance fluctuations that might be more closely related to transient stress.

The secondary findings are consistent with heterogeneous literature on activity and stress. In college students, Felez-Nobrega et al. [14] reported associations of perceived stress with self-reported MVPA and selected sedentary behaviors, whereas corresponding associations were less evident for monitor-derived measures.

Accelerometers quantify movement intensity but not context, which may matter for perceived stress. Similarly, our sleep measure represented diary-recorded bed-to-wake time rather than sleep quality or objectively measured sleep duration; self-reported and actigraphy-derived sleep duration show only moderate agreement [15].

Strengths include 28-day free-living accelerometry, an absolute minimum daily wear-time criterion with at least 14 valid days, individually measured RMR, separate physical and psychological stress assessments, baseline-adjusted models, and sensitivity analyses. Participants contributed a mean of more than 22 valid days, providing relatively dense characterization of habitual activity.

Limitations include the small, male-only sample; sample reduction caused by accelerometer data-quality requirements; questionnaire-based TEI; estimated rather than criterion-measured TEE; and assignment of RMR to periods without accelerometer-derived intensity, which may introduce error when actual expenditure differs from RMR. Sleep was self-reported, stress was assessed only twice, and academic, social, and health-related confounding was not comprehensively measured. Wide CIs indicate limited precision and do not exclude moderate associations. The findings may not generalize to women, non-student populations, or populations with different dietary and activity patterns, and the observational design precludes causal inference.

In conclusion, no clear association between estimated free-living energy balance and subsequent physical or psychological stress was detected in this exploratory sample. These preliminary effect estimates can inform larger studies using repeated stress assessments and more temporally aligned measures of energy intake, expenditure, and sleep.

## Data Availability

All data produced in the present study are available upon reasonable request to the authors.

## Declarations

### Ethical approval and consent to participat

This study was conducted in accordance with the principles of the Declaration of Helsinki and was approved by the Ethics Review Committee of Kyushu Sangyo University (approval no. 2020-0006). All participants received verbal and written explanations regarding the study procedures and the handling of their personal information. Written informed consent to participate was obtained from all participants.

### Competing interests

The author declares no competing interests.

### Funding

This work was supported by JSPS KAKENHI Grant Number JP20K19662.

### Data availability

The data supporting the findings of this study are available from the corresponding author upon reasonable request. The data are not publicly available because they contain information from human participants and their sharing is subject to ethical and privacy considerations.

## Acknowledgements

The author thanks the participants for their involvement in the study.

## Notes

### Competing Interest Statement

The authors have declared no competing interest.

### Author Declarations

Ethics committee of Kyushu Sangyo University gave ethical approval for this work.

## References

1. Mountjoy M, Ackerman KE, Bailey DM, Burke LM, Constantini N, Hackney AC, Heikura IA, Melin A, Pensgaard AM, Stellingwerff T et al: 2023 International Olympic Committee’s (IOC) consensus statement on Relative Energy Deficiency in Sport (REDs). British Journal of Sports Medicine 2023, 57(17):1073–1098.

2. Pensgaard AM, Sundgot-Borgen J, Edwards C, Jacobsen AU, Mountjoy M: Intersection of mental health issues and Relative Energy Deficiency in Sport (REDs): a narrative review by a subgroup of the IOC consensus on REDs. British Journal of Sports Medicine 2023, 57(17):1127–1135.

3. Wunsch K, Fiedler J, Bachert P, Woll A: The Tridirectional Relationship among Physical Activity, Stress, and Academic Performance in University Students: A Systematic Review and Meta-Analysis. International Journal of Environmental Research and Public Health 2021, 18(2):739.

4. Gardani M, Bradford DRR, Russell K, Allan S, Beattie L, Ellis JG, Akram U: A systematic review and meta-analysis of poor sleep, insomnia symptoms and stress in undergraduate students. Sleep Medicine Reviews 2022, 61:101565.

5. Sato H, Nakamura H, Nishida Y, Shirahata T, Yogi S, Akagami T, Soma M, Inoue K, Niitsu M, Mio T et al: Energy expenditure and physical activity in COPD by doubly labelled water method and an accelerometer. ERJ Open Research 2021, 7(2):00407–02020.

6. Weir JBdV: New methods for calculating metabolic rate with special reference to protein metabolism. The Journal of Physiology 1949, 109(1-2):1–9.

7. Kobayashi S, Murakami K, Sasaki S, Okubo H, Hirota N, Notsu A, Fukui M, Date C: Comparison of relative validity of food group intakes estimated by comprehensive and brief-type self-administered diet history questionnaires against 16 d dietary records in Japanese adults. Public Health Nutrition 2011, 14(7):1200–1211.

8. Kobayashi S, Honda S, Murakami K, Sasaki S, Okubo H, Hirota N, Notsu A, Fukui M, Date C: Both Comprehensive and Brief Self-Administered Diet History Questionnaires Satisfactorily Rank Nutrient Intakes in Japanese Adults. Journal of Epidemiology 2012, 22(2):151–159.

9. Murakami H, Kawakami R, Nakae S, Yamada Y, Nakata Y, Ohkawara K, Sasai H, Ishikawa-Takata K, Tanaka S, Miyachi M: Accuracy of 12 Wearable Devices for Estimating Physical Activity Energy Expenditure Using a Metabolic Chamber and the Doubly Labeled Water Method: Validation Study. JMIR Mhealth Uhealth 2019, 7(8):e13938.

10. Yoshida A, Ishikawa-Takata K, Tanaka S, Suzuki N, Nakae S, Murata H, Taguchi M, Higuchi M: Validity of Combination Use of Activity Record and Accelerometry to Measure Free-Living Total Energy Expenditure in Female Endurance Runners. Journal of Strength & Conditioning Research 2019, 33(11):2962–2970.

11. Okubo H, Sasaki S, Rafamantanantsoa HH, Ishikawa-Takata K, Okazaki H, Tabata I: Validation of self-reported energy intake by a self-administered diet history questionnaire using the doubly labeled water method in 140 Japanese adults. European Journal of Clinical Nutrition 2008, 62(11):1343–1350.

12. Nagayoshi S, Oshima Y, Ando T, Aoyama T, Nakae S, Usui C, Kumagai S, Tanaka S: Validity of estimating physical activity intensity using a triaxial accelerometer in healthy adults and older adults. BMJ Open Sport & Exercise Medicine 2019, 5(1):e000592.

13. Lesage F-X, Berjot S, Deschamps F: Clinical stress assessment using a visual analogue scale. Occupational Medicine 2012, 62(8):600–605.

14. Felez-Nobrega M, Bort-Roig J, Briones L, Sanchez-Niubo A, Koyanagi A, Puigoriol E, Puig-Ribera A: Self-reported and activPALTM-monitored physical activity and sedentary behaviour in college students: Not all sitting behaviours are linked to perceived stress and anxiety. Journal of Sports Sciences 2020, 38(13):1566–1574.

15. Lauderdale D, Knutson K, Yan L, Liu K, Rathouz P: Self-Reported and Measured Sleep Duration: How Similar Are They? Epidemiology 2008, 19(5):838–845.

